# One Health Longitudinal Study Protocol on Zoonotic and Vector-Borne Diseases in Battambang province, Cambodia: An Inter- Sectoral Approach

**DOI:** 10.64898/2026.03.14.26347916

**Authors:** Andrea Antoniolli, Mallorie Hide, Hélène Guis, Vincent Herbreteau, Sébastien Boyer, Sokleaph Cheng, Kunthy Nguon, Sopheak Sorn, Julia Guillebaud, Juliana Aizawa Porto De Abreu, Savatey Hak, Kimyeun Oeurn, Kimsear Nov, Janin Nouhin, Vibol Hul, Veasna Duong, Erik A Karlsson, Sopheak Chhay, Phanith Kong, Florian Commans, Chanchakriya Sam, Pavly Gov, Vathanak Heng, Kimsan Souv, Sitha Sin, Téphanie Sieng, Tineke Cantaert, Bertrand Guillard, Simon Cauchemez, Sidonn Krang, Flavie Goutard, Sowath Ly, Anne-Laure Bañuls, Claude Flamand

## Abstract

**Background:** Tropical low- and middle-income countries are highly vulnerable to zoonoses and vector-borne diseases, with risks amplified by climatic events, environmental change, and limited surveillance capacity. Cambodia is particularly exposed due to its ecological diversity, seasonal flooding, and rapidly changing land use. Globally, however, field based One Health approaches remain under-implemented, limiting practical evidence on how to address these complex threats.

**Methods:** This protocol describes a longitudinal One Health study conducted in three villages of Battambang province, Cambodia, designed to investigate the prevalence and transmission dynamics of zoonotic and potentially zoonotic pathogens at the human–animal–environment interface. The study examines how vector density, diversity, and pathogen circulation are influenced by hydrological variation and seasonality, and assesses the socio-demographic, behavioral, and environmental factors shaping transmission. Integrated data will be collected through serological and molecular analyses in humans and animals, environmental sampling, and entomological surveillance, enabling cross-compartmental and spatiotemporal analyses.

**Expected Results:** The study will generate integrated, cross-sectoral data to characterize pathogen exposure patterns, identify high-risk populations and practices, and inform targeted public health, veterinary, and environmental interventions.

**Conclusions:** By sharing this protocol, the work addresses a global methodological gap in operationalizing One Health in the field and supports the development of integrated surveillance strategies in climate-sensitive, resource-limited settings.

## Introduction

Emerging and re-emerging zoonotic and vector-borne diseases are a major public health concern in low- and middle-income countries, particularly in tropical regions such as Cambodia. Rapid urbanization, deforestation, agricultural expansion, and climate variability, leading to environmental changes such as flooding and drought, are altering vector habitats and pathogen dynamics. These shifts, combined with increasing human-animal-environment interactions, create conditions conducive to pathogen spillover and sustained transmission [1]. Under-resourced surveillance systems across humans, animal and environmental sectors, limited diagnosis capacity and the limited adoption of coordinated multisectoral analytical approaches further hinder the accurate assessment of disease burden and transmission dynamics. This gap not only reduces the understanding but also reduces the effectiveness of integrated, public health, veterinary and environmental interventions [2].

Located within the Indo-Burma biodiversity hotspot, Cambodia is particularly prone to ecological and anthropogenic pressures that increase the risks of infectious disease emergence [3]. Hydroclimatic phenomena, such as flooding and shifting precipitation patterns, compound these challenges by influencing pathogen survival, vector distribution, wildlife movement, and the spread of diseases through socio-ecosystems [4].

Globalization, through trade, tourism, and human, animal or product movement, adds another layer of complexity, enabling the introduction and rapid dissemination of zoonotic and vector-borne pathogens [3,4]. Cambodia’s epidemiological landscape is marked by a high burden of zoonotic and potentially-zoonotic diseases like avian influenza, leptospirosis, melioidosis [6–10], and vector-borne diseases, such as dengue, chikungunya and Japanese encephalitis [11–20]. These diseases affect both human and animal populations alike, with consequences for health, livelihoods, food security and ecosystem stability. The country’s dependence on agriculture–sustaining over half of all households, with a majority raising poultry or livestock [21] – intensifies these risks, by bringing humans, domestic animals and wildlife into close and prolonged contact, while also increasing the vulnerability of rural communities to livestock morbidity, mortality and reduced productivity [22].

The One Health framework, which holistically integrates human, animal and environmental health is widely recognized as essential to tackling these interconnected challenges [23–25]. However, the true One Health implementation often remains fragmented, and comprehensive field-based One Health studies are still rare [26,27] limiting our understanding of the drivers of transmission and the opportunities for coordinated, cross-sectoral intervention [28,29].

This protocol describes the design of a longitudinal One Health study conducted in Battambang province, located in the western part of Cambodia, as part of PREACTS-AFRICAM study, which constitutes the first phase of the PREZODE (Preventing Zoonotic Disease Emergence) initiative. The overall study aims to investigate zoonotic and arboviral pathogen transmission dynamics in rural communities, taking into account climatic and environmental factors, livestock and wildlife interactions and human behavioral drivers. The goal is to generate actionable insights that support risk mitigation, guide targeted interventions across human, animal and environmental sectors and strengthen integrated surveillance systems in Cambodia. The primary objective is to measure the prevalence of target pathogens and identify transmission pathways across the three One Health compartments. Secondary objectives include assessing risk factors at individual, household and community levels, and providing evidence to guide targeted public health interventions.

## Methods

### Aim

This study aims to improve the understanding of zoonotic and vector-borne disease dynamics across humans, animals, and the environment in rural Cambodia, accounting for hydrological variations and seasonality. The focus is on identifying disease prevalences, transmission drivers and risk factors to inform prevention and control strategies for emerging and re-emerging infectious diseases.

### Study design and setting

This is a longitudinal, prospective cohort study conducted over 21 months in three rural villages in Ek Phnom district, Battambang province, Cambodia (Fig 1). Battambang province, located along the Tonle Sap Lake, has a population of approximately 1.3 million. Its provincial capital, Battambang town, lies about 290 km from the national capital, Phnom Penh. The three study villages are situated along the 250 km-long Sangker River: Prek Norin, a semi-rural village located 20 km from the provincial capital; Prek Trab, a rural village situated 30 km away; and Bak Prea, a seasonal “floating village” that is flooded for several months during the rainy season, located 50 km away from the provincial capital with limited road access.

**Fig 1:**
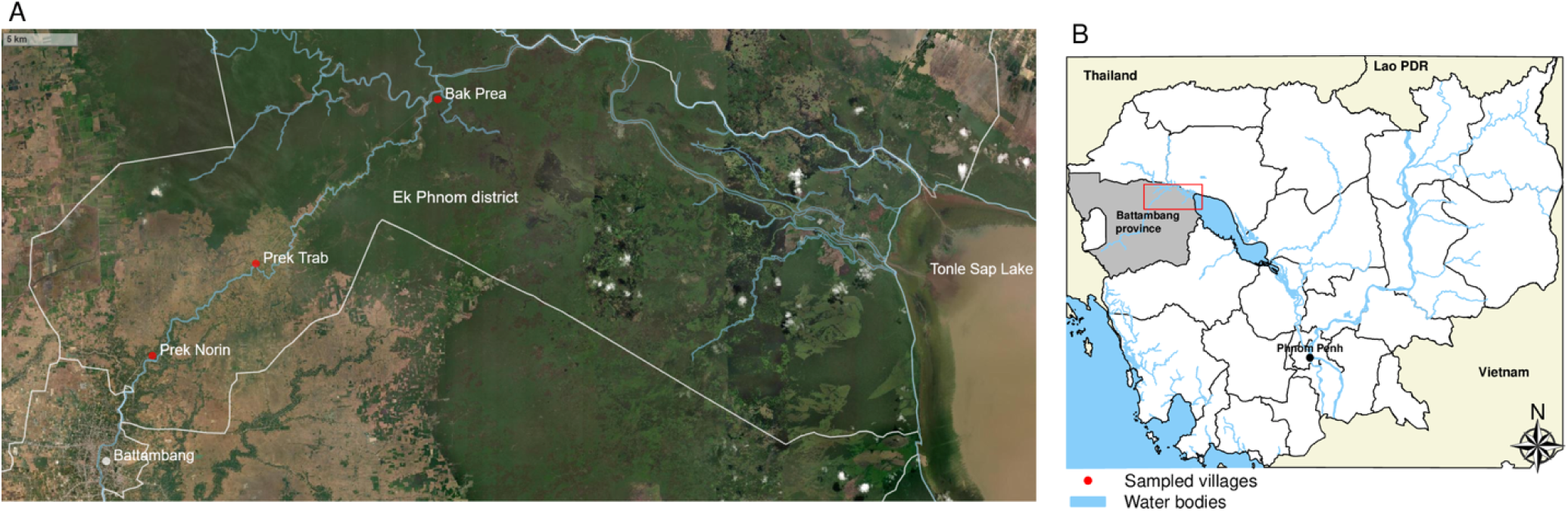
Study area and location of the three selected villages (Prek Norin, Prek Trab and Bak Prea, Battambang province). (A) Study Zone. https://leafletjs.com/ Leaflet | Tiles © Esri – Source: Esri, i-cubed, USDA, USGS, AEX, GeoEye, Getmapping, Aerogrid, IGN, IGP, UPR-EGP, and the GIS User Community: open-source JavaScript library, BSD 2-Clause Licence. Map generated using R version 4.3.1. (B) Cambodia. All materials are shapefiles under the Creative Commons Attribution (CC BY) licence from https://data.opendevelopmentcambodia.net, https://data.opendevelopmentmekong.net, and https://data.humdata.org. Map generated using R version 4.3.1.

The study integrates human, animal, vector and environmental surveillance within a One Health framework. Participants are enrolled during an initial recruitment campaign (M0) and followed every six months across four data collection rounds (M0, M6, M12, M18) using a fixed cohort design, with no new enrolments after the baseline. In parallel, concurrent sampling of domestic animals (cattle, swine, poultry), wildlife (rodents), water and soil is conducted to support integrated One Health surveillance. Mosquito collection is conducted more frequently, every 3 months over eight rounds. Entomological surveys are staggered to begin one month before each human sampling round to account for both extrinsic and intrinsic incubation periods of vector-borne diseases.

### Sampling strategy

Households are stratified using a random sampling approach based on animal ownership, targeting 450 households (150 per village) with an estimated 1,350 human participants. Prior to the baseline survey, a preparatory mission with village chiefs records the current population census of each study village, the number and types of animals in each household. These data inform species-specific sampling targets per village based on the presence of animals per household.

The animal-owning stratum is defined in three steps:

1. All households owning pigs or cattle are included, due to their low numbers and relevance as potential zoonotic reservoirs.
2. Among the remaining households, 30% of those owning poultry (chickens or ducks) are randomly selected.
3. Additional households without animals are randomly selected to reach the target of 150 households per village.

To account for refusals or ineligibility, 20 backup households per stratum are randomly selected from the remaining eligible pool. The longitudinal surveillance will be conducted at the household level for humans, and more specifically at the herd level for domestic animals, meaning that the same households will be followed throughout the duration of the study, even if the individual livestock animals are not the same.

### Domestic animal sampling

Among animal-owning households selected for human sampling, 75 households (25 per village) are randomly selected for biological sampling of domestic animals. The number of animals sampled per species and per village is determined based on the preparatory census to ensure adequate coverage of major livestock species for serological and microbiological analyses. All households with pigs are included due to their very small number. Random sampling is then performed among households with cattle. If the total remains below 25, additional households with poultry only are selected. To address potential refusals or cases of inaccessibility, 10 backup households per village are selected.

### Rodent sampling

Rodent trapping is conducted in a 90-household subsample (30 per village) from the enrolled households. In each village, the 25 households where domestic animals are sampled, along with 5 additional households without animals are randomly selected. A stratified sampling strategy is applied with an allocation ratio of 5:1 between households with and without livestock animals.

To address potential refusals or cases of inaccessibility, 10 backup households per village are selected. An additional 40 rodent traps are deployed per village.

### Environmental sampling

Environmental sampling is conducted in the same 30 households as rodent trapping, (including the 25 households used for domestic animal sampling and 5 households without animals) targeting both peri-domestic and rural areas for bacteriological and parasitological detection. A total of 10 backup households per village are selected. Additionally, 45 samples per village are collected.

### Vector sampling

Vector sampling is conducted in a sub-sample of 75 households (25 per village) selected within the human cohort. To ensure diverse exposure contexts and sufficient power for comparative analyses, an approximate 1:1 ratio is targeted, between households with target animal species and those without any animal. All households with pigs are automatically included due to their relevance as amplifying hosts of Japanese encephalitis, [30] while the remaining households are randomly selected to maintain balance. Additionally, 10 backup households per village (five per stratum) are identified.

### Inclusion criteria

Participants must meet the following conditions to be eligible for enrollment:

- Be between 2 and 75 years of age;
- Have resided in one of the study villages (Prek Norin, Prek Trab, or Bak Prea) for at least six months prior to recruitment and plan to remain throughout the study period;
- Provide written informed consent (for adults);
- For participants aged 2–17 years, informed consent must be obtained from one parent or guardian, along with the child’s oral assent for those aged 13-17 years old

### Non-Inclusion criteria

Individuals will not be eligible for participation if they:

- Are unable to understand the study procedures or provide informed consent;
- Are under legal guardianship or deprived of liberty by court order;
- Have medical conditions that preclude safe participation in interviews or biological sampling;
- Decline to participate in the study.

### Intervention

This is a longitudinal observational study with no planned medical or behavioural interventions. The focus is on the collection of data and biological samples across human, animal, and environmental compartments for surveillance and research purposes.

### Sample size

Given the multitude of pathogens and the lack of prior knowledge regarding actual seroprevalence, a 50% seroprevalence for a specific pathogen is assumed to apply the most conservative approach and ensure a sufficient sample size.[31] With a 95% confidence level and a design effect of 2, a total sample size of 1,350 individuals will yield a margin error of 0.04 for prevalence estimates across the entire study area. At the village level (450 individuals per village), the margin error is 0.07. Based on an average of 3 people investigated per household, 150 households are randomly selected per village, giving a total of 450 households across the three study villages.

For animal sampling, assuming a 50% prevalence for a specific pathogen, a 95% confidence level and a design effect of 2, a total of 1,500 animals will provide a margin error of 0.03, with 1500 animals for prevalence estimates across the entire study area, and 0.06 at the village level (500 animals per village, 125 cattle, 125 swine and 250 poultry).

## Material collection

### Humans

From each enrolled participant, a venous blood sample (up to 10 mL for individuals aged >7 years, 5 mL for children under 7 years) will be collected at each round for serological analyses.

When participants present with fever or other symptoms suggestive of infection during a visit, they will be promptly referred to the nearest healthcare facility for appropriate clinical care and additional ad hoc diagnostic testing, in accordance with national guidelines.

### Domestic animals

In household owning animals, up to 10 poultry will be sampled, with one oral swab and one rectal swab collected per bird (maximum 20 samples per household). For cattle and pigs, up to 5 individuals per species will be sampled, with oral and rectal swabs, and blood and urine when possible (maximum 20 samples per household). This yields a theoretical maximum of 1,500 samples per village, 4,500 per round across the three villages), and 18,000 over the study period. (Table 1). Whenever possible, the same individual domestic animals will be resampled during follow-up visits using identifying features (photos, names, physical marks). This will not be feasible for poultry, given their relatively short lifespan. Sample collections will follow standard practices and will be conducted with the animal owners’ consent and cooperation. Veterinarians from the Directorate of Animal Health and Production (GDAPH) of the Ministry of Agriculture will oversee and supervise all animal sampling, ensuring it’s conducted by qualified professionals in line with ethical standards and animal-welfare guidelines. GDAHP officers will be present at all field activities and will be responsible for the supervision of animal sampling. Sample collection will follow standardized procedures to ensure efficient collection with minimal impact on animal welfare.

**Table 1:**
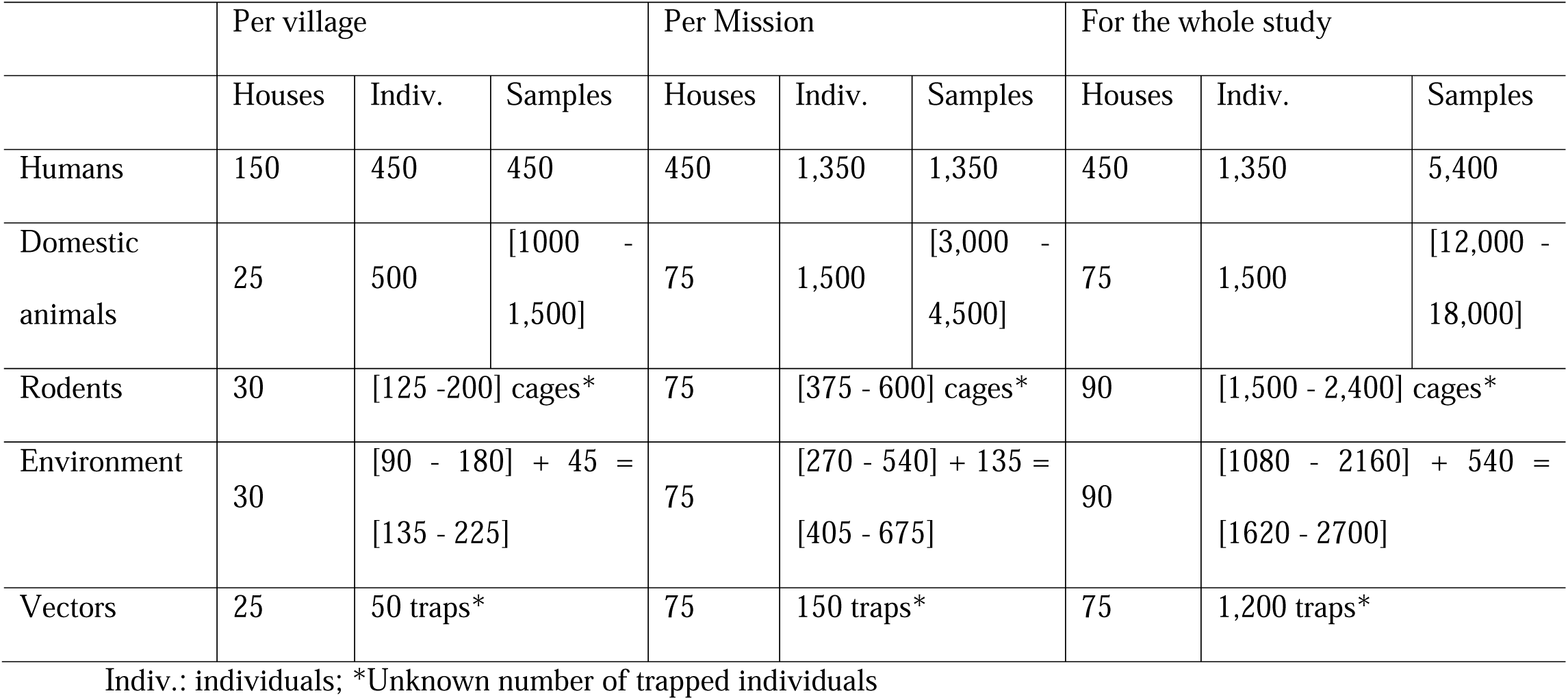
Sample design of the longitudinal study by village and mission.

|  | Per village |  |  | Per Mission |  |  | For the whole study |  |  |
| --- | --- | --- | --- | --- | --- | --- | --- | --- | --- |
|  | Houses | Indiv. | Samples | Houses | Indiv. | Samples | Houses | Indiv. | Samples |
| Humans | 150 | 450 | 450 | 450 | 1,350 | 1,350 | 450 | 1,350 | 5,400 |
| Domestic animals | 25 | 500 | [1000 - 1,500] | 75 | 1,500 | [3,000 - 4,500] | 75 | 1,500 | [12,000 - 18,000] |
| Rodents | 30 | [125 -200] cages* |  | 75 | [375 - 600] cages* |  | 90 | [1,500 - 2,400] cages* |  |
| Environment | 30 | [90 - 180] + 45 =<br>[135 - 225] |  | 75 | [270 - 540] + 135 =<br>[405 - 675] |  | 90 | [1080 - 2160] + 540 =<br>[1620 - 2700] |  |
| Vectors | 25 | 50 traps* |  | 75 | 150 traps* |  | 75 | 1,200 traps* |  |

Indiv.: individuals; \*Unknown number of trapped individuals

### Rodents

Rodent trapping will be performed using 3 to 5 live-capture cages per household, depending on logistical constraints and dwelling size. Trapping is repeated, inside and around each household, for two consecutive nights per mission. An additional 40 traps per village will be placed in ecologically relevant habitats (e.g. rice fields, floodplains, drylands, fruit tree orchards, shrub-covered fallow lands, and wooded areas with natural forests or plantation crops), to capture a diversity of species. This results in 310 to 390 traps deployed per round across the three villages. Non-target or protected species will be released immediately and unharmed at the capture site when traps are opened. Rodents will be captured using individual live traps and will not be retained for extended periods. Target rodents will be processed immediately after capture and humanely euthanized by isoflurane inhalation. After transportation, kidney tissues will then be collected and tested in accordance with the Guide for the Care and Use of Laboratory Animals of the National Institutes of Health.[32] All sampling procedures will be performed under supervision of trained personnel from the Forestry Administration of the Ministry of Agriculture, Forestry and Fisheries (MAFF), in full compliance with ethical and animal welfare guidelines.

### Environment

Environmental sampling will be performed, collecting two water samples (drinking and wastewater, 50mL each) and one surface soil sample (250mg) per household per mission. When available, samples related to animal presence (wastewater, drinking water and soil) will be collected with a maximum of six environmental samples per household. Additionally, 45 samples per village will be collected from several rural sites (e.g., rice fields, grazing paths, riverbanks). At each site, one surface water sample (50ml) and four soil samples (250mg each) will be collected at depths of 1, 30, 60 and 90cm. In total, up to 675 environmental samples will be collected per mission (540 household-based, 133 site-based), for 2,700 over the study.

In total, up to 675 environmental samples per mission (540 household-based, 135 site-based), for 2,700 over the study.

### Vectors

In selected household, one BG-Sentinel trap and one CDC light trap will be deployed outdoors for 24 hours [33], resulting in 150 traps per mission (2 traps × 75 households). This approach allows for temporal and spatial comparisons of vector abundance and species composition across different ecological and domestic environments and facilitates linkage with human and animal infection or immune status. Ectoparasites collected from trapped rodents will also be preserved and analyzed to broaden vector surveillance.

### Survey status and timeline

As of the submission date (August 2025), participant recruitment has begun but is not complete, data collection is ongoing, and no study outcomes have been analyzed or reported. In January 2025, a preliminary visit was conducted in the three study villages with the village chiefs to compile an exhaustive household list based on a rapid census of residents and targeted animal species; these preparatory activities established the sampling frame and did not generate study outcome data. Participant enrollment began on 24 June 2025 and is anticipated to be completed by 30 August 2025. Data collection, including participant follow-up, will continue for 18 months and is expected to be completed by 31 December 2026. The entomological component (vector sampling) will run from 2 June 2025 to 31 March 2027. The overall schedule of multi-compartment sampling missions is shown in Fig 2. Final integrated analyses are expected by September 2027, i.e., within ∼6 months after completion of the last data collection activity (entomological sampling in March 2027).

**Fig 2:**
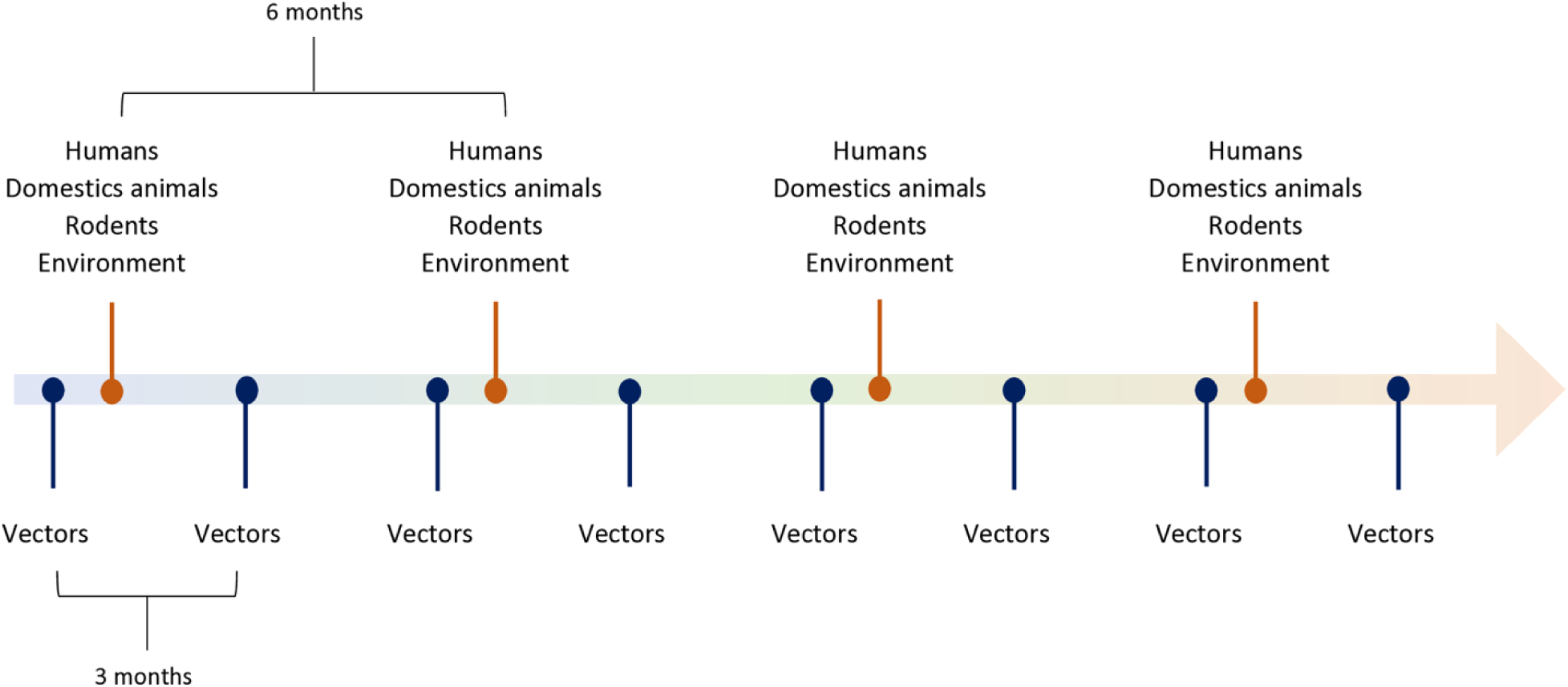
Timeline of mosquito, human, domestic animal, rodent, and environmental sampling missions over the course of the longitudinal study.

## Outcomes

### Primary outcomes

We will explore exposure–in humans–or presence/exposure–in animals and in the environment–of 19 pathogens, including nine bacteria (*Leptospira interrogans, Burkholderia pseudomallei, Orientia tsutsugamushi, Rickettsia typhi, Coxiella burnetii, Borrelia burgdorferi s.l* and 3 enterobacteria: *Salmonella* Typhi*, Salmonella* Paratyphi*, Shigella spp.*), five parasites (*Toxoplasma gondii, Entamoeba histolytica, Schistosoma japonicum* group*., Clonorchis sinensis, Opisthorchis viverrini*) and seven viruses (5 arboviruses: dengue (DENV), zika (ZIKV), chikungunya (CHIKV), Japanese encephalitis (JEV), west Nile (WNV); orthohantaviruses and avian influenza).

Table 2 details the pathogens investigated in humans by serology, in domestic animals, rodents and the environment by RT-qPCR.

**Table 2:** Overview of Investigated Pathogens.

|  | Pathogen | Human exposure<br>(seroprevalence) | Domestic<br>animal<br>infection<br>(qPCR) | Rodent<br>infection<br>(qPCR) | Environmental<br>presence<br>(qPCR) |
| --- | --- | --- | --- | --- | --- |
| Bacteria | *Leptospira interrogans* | ? | ? | ? | ? |
|  | *Burkholderia pseudomallei* | ? | ? | ? | ? |
|  | *Orientia tsutsugamushi* |  |  | ? | ? |
|  | *Rickettsia typhi* |  |  | ? | ? |
|  | *Coxiella burnetii* | ? | ? | ? | ? |
|  | *Borrelia burgdorferi s.l* | ? | ? | ? | ? |
|  | Enterobacteria |  | ? | ? | ? |
| Parasites | *Toxoplasma gondii* | ? | ? | ? | ? |
|  | *Entamoeba histolytica* |  | ? | ? | ? |
|  | *Schistosoma japonicum*<br>group |  | ? | ? | ? |
|  | *Clonorchis sinensis* |  | ? |  | ? |
|  | *Opisthorchis viverrini* |  | ? |  | ? |
| Viruses | Arboviruses (DENV,<br>ZIKV, CHIKV, JEV,<br>WNV) | ? |  |  |  |
|  | Hantaviruses | ? |  | ? |  |
|  | Avian influenza | ? | ? |  |  |

### Secondary outcomes

Secondary outcomes include the detection of mosquito salivary biomarkers in human samples to characterize exposure to mosquito bites[13,34]. We also aim to identify environmental, socio-economic and behavioral risk factors associated with exposure to these pathogens in both human and animals based on data collected through questionnaires. Additionally, we will assess the spatial and temporal distribution of identified zoonotic pathogens using molecular data to map their circulation within the study sites.

### Predictors

Data collected through structured questionnaires and satellite-derived environmental variables serve as predictors to assess seroprevalence in humans and positivity rates in animal and environmental samples. Additionally, molecular positivity in animals and environmental samples will be analyzed as a predictor of human seroprevalence and to investigate potential transmission pathways across compartments.

### Sample processing and biological analysis

#### Pathogen detection in biological samples

Blood tubes will be transported to the Virology Unit, and Medical Biology Laboratory of Institut Pasteur du Cambodge (IPC) as soon as possible within 24 hours after collection, and serum samples will be cryopreserved at-80°C. For each biological sample collected, the date of collection and the time of arrival at IPC laboratory will be recorded. (Table 3)

**Table 3:** Type of sample and Sampling processing.

| Sample | Analysis | Volume (ml) | Sampling Method | Sample processing |
| --- | --- | --- | --- | --- |
| Human blood samples | Serology | 10ml (>7yo)/<br>5ml (≤7yo blood (serum) | Dry tubes | *Sample collected during the interview after obtaining the signature of the informed consent: Immediately shipped to IPC laboratory at temperature at 4-8°C. Serum preparation and cryopreservation (-80°C) for storage before analyses.* |
| Domestic animals (poultry, swine, cattle) oral and rectal swabs and blood and urine samples | Molecular detection |  |  | *Animal sampling during field investigations: On-site cryopreservation in liquid nitrogen, followed by transfer to the Virology Unit/MBL (IPC), for storage at -80°C before analyses.* |
| Rodent (rodent) organs and blood samples | Molecular detection | Organs, blood, oral and rectal swabs | Dissection for organ collection<br>Storage in microtubes | *Animal sampling during field investigations: On-site cryopreservation in liquid nitrogen, followed by transfer to the Virology Unit/MBL (IPC), for storage at -80°C before analyses.* |
| Environmental samples | Molecular detection | Soil (250mg)<br>Water (50ml) | Auger<br>Swinnex filtration | *Environmental sampling during field investigations: On-site cryopreservation in liquid nitrogen, followed by transfer to the MBL (IPC), for storage at -80°C before analyses.* |

To avoid multiple freeze-thaw cycles, samples will be aliquoted prior to freezing. Animal samples will be cryopreserved on-site at-80°C and then be stored in a dedicated buffer. Environmental samples (water and soil) will be collected in dry microtubes and stored in liquid nitrogen until DNA extraction.

Serological analyses of human samples will be performed using a bead-based multiplex microsphere immunoassay (MIA)[35–38] and standard serological assays (ELISA and MAT). Established neutralization tests are used to facilitate and validate the interpretation of arbovirus results. Bead preparation for the Luminex MIA will be conducted at IPC following a validated protocol for five arboviruses. This Luminex MIA has been benchmarked against PRNT assays using 22 antigenic targets, including virus-like particles, NS1, and domain III of the envelope protein of the four dengue serotypes, Zika, West Nile, chikungunya (envelope protein), and JEV (NS1 and envelope protein). Additional pathogens from the targeted pathogen list may be included in the multiplex serological panel as further assay developments become available. (Table 4)

**Table 4:**
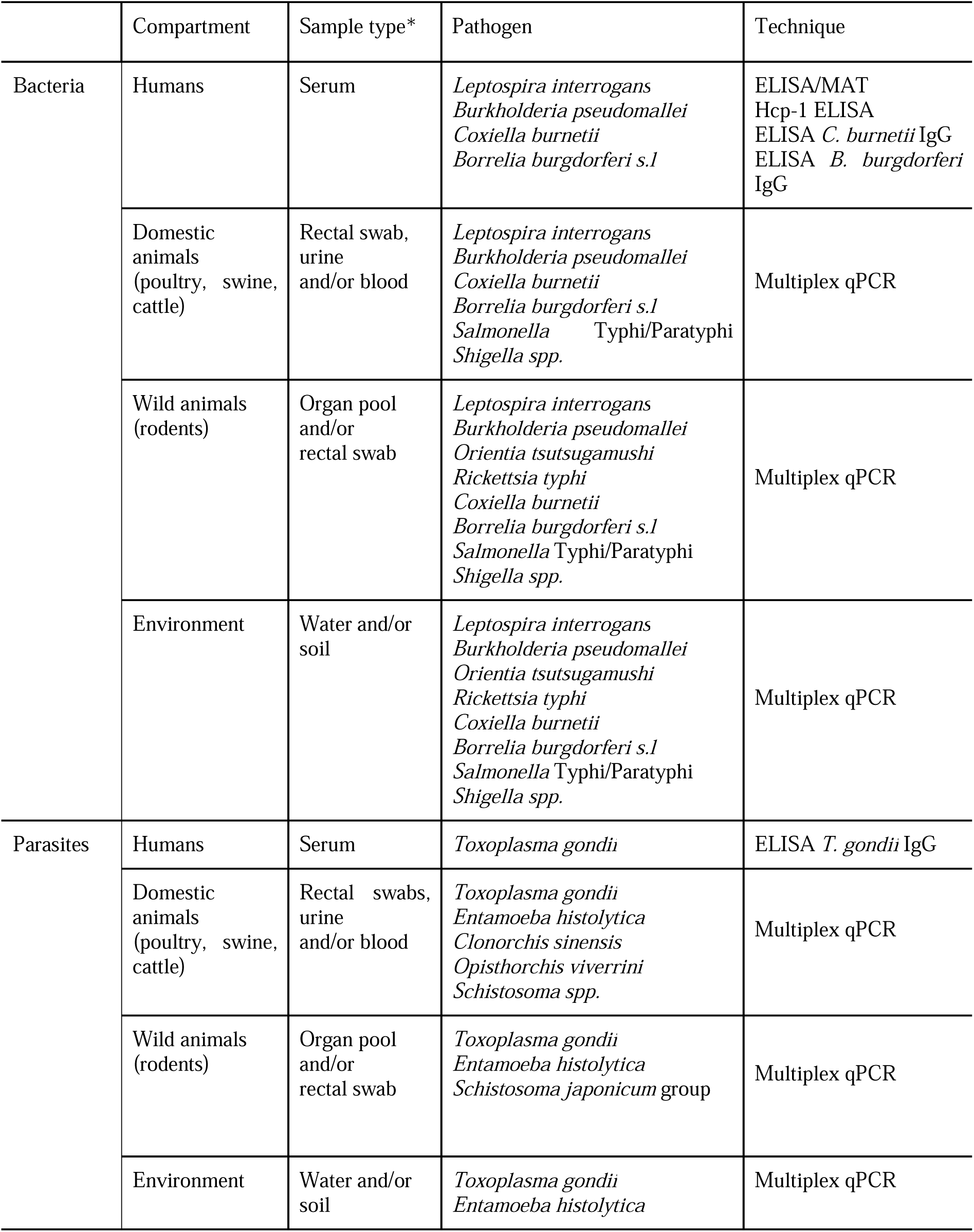
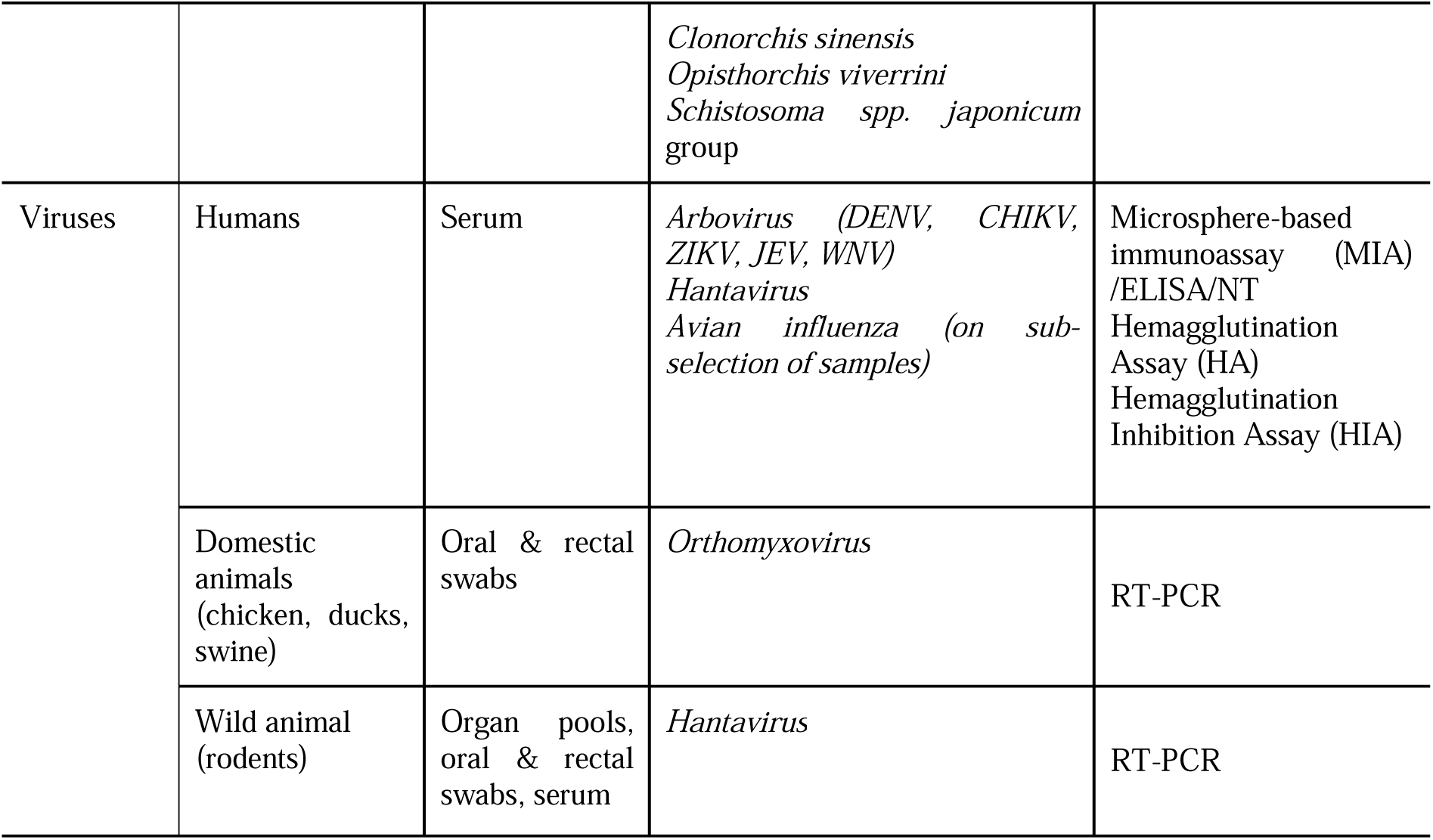
Type of Pathogens and Biological samples investigated.

|  | Compartment | Sample type* | Pathogen | Technique |
| --- | --- | --- | --- | --- |
| Bacteria | Humans | Serum | *Leptospira interrogans*<br>*Burkholderia pseudomallei*<br>*Coxiella burnetii*<br>*Borrelia burgdorferi s.l* | ELISA/MAT<br>Hcp-1 ELISA<br>ELISA *C. burnetii* IgG<br>ELISA *B. burgdorferi* IgG |
|  | Domestic animals (poultry, swine, cattle) | Rectal swab, urine and/or blood | *Leptospira interrogans*<br>*Burkholderia pseudomallei*<br>*Coxiella burnetii*<br>*Borrelia burgdorferi s.l*<br>*Salmonella* Typhi/Paratyphi<br>*Shigella spp.* | Multiplex qPCR |
|  | Wild animals (rodents) | Organ pool and/or rectal swab | *Leptospira interrogans*<br>*Burkholderia pseudomallei*<br>*Orientia tsutsugamushi*<br>*Rickettsia typhi*<br>*Coxiella burnetii*<br>*Borrelia burgdorferi s.l*<br>*Salmonella* Typhi/Paratyphi<br>*Shigella spp.* | Multiplex qPCR |
|  | Environment | Water and/or soil | *Leptospira interrogans*<br>*Burkholderia pseudomallei*<br>*Orientia tsutsugamushi*<br>*Rickettsia typhi*<br>*Coxiella burnetii*<br>*Borrelia burgdorferi s.l*<br>*Salmonella* Typhi/Paratyphi<br>*Shigella spp.* | Multiplex qPCR |
| Parasites | Humans | Serum | *Toxoplasma gondii* | ELISA *T. gondii* IgG |
|  | Domestic animals (poultry, swine, cattle) | Rectal swabs, urine and/or blood | *Toxoplasma gondii*<br>*Entamoeba histolytica*<br>*Clonorchis sinensis*<br>*Opisthorchis viverrini*<br>*Schistosoma spp.* | Multiplex qPCR |
|  | Wild animals (rodents) | Organ pool and/or rectal swab | *Toxoplasma gondii*<br>*Entamoeba histolytica*<br>*Schistosoma japonicum* group | Multiplex qPCR |
|  | Environment | Water and/or soil | *Toxoplasma gondii*<br>*Entamoeba histolytica* | Multiplex qPCR |
|  |  |  | *Clonorchis sinensis*<br>*Opisthorchis viverrini*<br>*Schistosoma* spp. *japonicum*<br>group |  |
| Viruses | Humans | Serum | *Arbovirus* (*DENV*, *CHIKV*,<br>*ZIKV*, *JEV*, *WNV*)<br>*Hantavirus*<br>*Avian influenza* (on sub-<br>selection of samples) | Microsphere-based<br>immunoassay (MIA)<br>/ELISA/NT<br>Hemagglutination<br>Assay (HA)<br>Hemagglutination<br>Inhibition Assay (HIA) |
|  | Domestic<br>animals<br>(chicken, ducks,<br>swine) | Oral & rectal<br>swabs | *Orthomyxovirus* | RT-PCR |
|  | Wild animal<br>(rodents) | Organ pools,<br>oral & rectal<br>swabs, serum | *Hantavirus* | RT-PCR |

The seroprevalence of *Leptospira interrogans* will be assessed using SERION ELISA kits for all samples and using the gold-standard microscopic agglutination test (MAT), employing 12 serovars relevant to Cambodia for a subsample of samples. The seroprevalence of *Burkholderia pseudomallei* is measured using the Hemolysin-coregulated protein 1 (Hcp-1) ELISA. For *Coxiella burnetii* (Phase 2, ESR1312G), *Borrelia burgdorferi s.l.* (ESR121G), and *Toxoplasma gondii* (ESR110G), seroprevalence is determined using dedicated SERION ELISA kits.

#### Mosquito salivary biomarkers

Mosquito salivary biomarkers will be analyzed in human serum samples to assess exposure to *Aedes* mosquito bites. Upon feeding, mosquitoes inject salivary proteins that can induce specific antibody responses in humans, serving as biomarkers of vector–host contact [34,39]. This serological approach, validated in various settings[13,34,40–43], enables direct quantification of exposure to mosquito bites using ELISA assays on collected serum samples.

### Data: collection, management analysis and integration

#### Data collection

Data will be collected using standardized questionnaires capturing socioeconomic, environmental, clinical, and household characteristics. Participant identifiers will be used to securely link survey data with biological, environmental, and entomological samples across all compartments. Socioeconomic data will include age, gender, occupation, household income, education, and travel history. Environmental data will cover housing type, household size, number of rooms, assets, and animal ownership. Detailed exposure to domestic mammals, poultry, fish, wild animals (including rodents) and to mosquitoes will be recorded. Clinical data will document vaccination status, reported symptoms and any history of infection with arboviruses, malaria, or other illnesses, to support the assessment of health patterns and risk factors. Knowledge, attitudes, and practices (KAP) related to disease awareness, healthcare access, and acceptance of public health interventions will also be collected. An animal questionnaire will be administered during each field mission to document species present, number of individuals, breeding purpose, rearing practices, contacts with other species, exposure to floodwater, and living conditions. Animal health indicators, symptom history, vaccination status, and treatments use will be recorded. Environmental samples will be collected to characterize potential pathogen reservoirs and contamination sources, including water characteristics (pH, conductivity, temperature), soil characteristics (Humidity, conductivity, temperature), and vegetation indices. Entomological surveys will gather data on vector species composition, and density through standardized trapping methods and laboratory analyses. All data will be recorded using electronic-questionnaire implemented in REDCap, ensuring standardized data collection, quality assurance, and traceability across study components.

#### Data management

All data will be stored on IPC’s password-protected institutional servers behind secure firewalls. Data requestors must store data within institutional infrastructures adhering to robust security standards. Data collected from human, animal, vector, and environmental compartments are systematically compiled and integrated into a centralized database. Questionnaire responses, serological and molecular diagnostic results, entomological data, and environmental indicators are linked using unique participant, household, and sampling site identifiers to enable cross-compartmental and longitudinal analyses.

#### Data analysis

Data analysis will include quality checks, removal of low-quality samples, normalization, imputation where appropriate and visualization using dimensionality reduction techniques. Primary analyses will apply standard epidemiological methods to assess seroprevalence across geographic areas, age groups, gender, habitat types, and risk factors such as animal ownership and pathogen presence in animals and the environment. Analyses will account for sampling design, clustering and post-stratification weights. Means and proportions will be compared across groups using parametric and non-parametric tests (e.g., Pearson, Spearman, Fisher). Univariate and multivariate regression models, relative risks and odds ratios will be used to identify associated factors. Mathematical models will be developed to reconstruct transmission dynamics using age-stratified serological data in humans. Additional models will explore the presence of pathogens in domestic animals, wildlife and environmental samples, integrating molecular detection data, human serological profiles and climatic variables. Entomological analyses will characterize vector diversity, abundance and infection rates across seasons and ecological contexts. Statistical models will assess associations between vector indices, and environmental or climatic conditions (e.g., rainfall, temperature, land use). The heterogeneity observed in entomological data will be compared to human salivary biomarkers specific to mosquito exposure, enabling the quantification of human-vector contact. These biomarkers will also be analyzed in relation to the seroprevalence and incidence of mosquito-borne pathogen infections, providing complementary insights into transmission intensity and individual exposure levels. Environmental and climate data will be integrated into spatio-temporal models to identify determinants of pathogen presence in the environment and infection risk patterns in human and animal populations. Remote sensing data and meteorological records will support the analysis of environmental factors such as flooding, vegetation cover, and habitat fragmentation, which will inform ecological risk mapping. Overall, these integrated analyses across compartments will provide a comprehensive understanding of transmission dynamics at the human-animal-environment interface, guiding the design of precision public health and veterinary interventions.

## Ethics and regulatory consideration

A written authorization from the Provincial Governor of Battambang (letter No. 2561-24, 19 September 2024) permits field access and inter-agency coordination. The protocol was approved by the Cambodian National Ethics Committee for Health Research (NECHR) on 17 March 2025 (Approval No. NECHR-120). On 11 June 2025, the study also received an oral endorsement from the Interministerial Coordinating Committee on One Health (IMCC-OH) following presentation and discussion of the protocol. IMCC-OH does not issue formal written approvals; its role is to coordinate and advise across sectors under the national One Health framework.

### Human participants

Written informed consent will be obtained from all adults; for minors, parental/guardian permission and age-appropriate assent will be secured. Venous blood volumes will not exceed 10 mL in individuals > 7 years and 5 mL in children ≤ 7 years per round. Participants presenting with fever or compatible symptoms at visits will be referred to the nearest healthcare facility for clinical care and any ad hoc testing, in line with national guidelines. Personal identifiers will be pseudonymized with restricted access to linkage files.

### Animal oversight and regulatory context

Cambodia does not have an institutional animal care and use committee (IACUC) system. In lieu of an IACUC, animal-sector activities are conducted under the authority, supervision, and active participation of the MAFF through its competent bodies: the GDAPH for livestock and domestic animals, and the Forestry Administration for wildlife (including small mammal trapping). Under prevailing national practice, no written permits are issued by MAFF for this type of work; instead, GDAPH and Forestry Administration officers attend fieldwork and supervise procedures on site to ensure adherence to animal-welfare standards and good practice. Field activities occur on private land or community spaces with prior permission from owners/authorities; no sampling is planned in protected areas. The study does not target protected species; any non-target or protected wildlife captured incidentally is immediately released unharmed at the capture site.

### Domestic animals (households)

With owner consent and cooperation, sampling includes oral and rectal swabs in poultry (up to 10 birds per household; 2 swabs/bird), and oral/rectal swabs with blood and urine when feasible in cattle and pigs (up to 5 individuals per species per household). Where possible, the same animals will be revisited at follow-up using identifying features (photos, names, physical marks). Procedures are minimally invasive and performed by trained veterinarians under GDAPH supervision, following standardized procedures to minimize distress and impact on animal welfare.

### Rodents (wildlife)

Live-capture traps (3–5 per household, two consecutive nights) will be set inside/around households, with ∼40 additional traps per village in ecologically relevant habitats (e.g., rice fields, orchards, shrub fallows, forest edges/plantations) to capture species diversity. Non-target or protected species are released immediately and unharmed upon trap opening. Target rodents are processed immediately after capture and humanely euthanized by isoflurane inhalation; kidney tissues are collected and tested following the Guide for the Care and Use of Laboratory Animals (NIH). All procedures are conducted under the supervision of trained Forestry Administration personnel of MAFF and comply with ethical and animal welfare guidelines.

Biosafety and data/sample management. Field teams use appropriate PPE and IPC/MAFF biosafety procedures. Samples are uniquely labeled, transported under cold chain, and stored in secure IPC facilities with controlled access. Logs and datasets are maintained in password-protected systems accessible only to authorized staff.

### Governance

The study operates within Cambodia’s One Health governance, with cross-sectoral coordination (MoH/NECHR, MAFF via GDAPH and the Forestry Administration and MoE). In the absence of a national IACUC mechanism, MAFF supervision and participation provide sectoral oversight for animal work, while NECHR approval covers human-subject procedures.

#### Consent

Eligible participants within selected households will be invited to participate in the study via an initial in-person interview. They will be informed of the characteristics and consequences of the study both verbally and in writing with the participant information sheet and consent form. If they agree to participate, they will sign the informed consent form and keep a copy. The investigator will also sign the consent form and keep a copy of the original. For minors aged 2-17 years old, consent will be obtained from one guardian or responsible adult along with an assent for age 13-17 years old. The consent form will be designed to allow individuals to agree to participate in this study and potential future research that might utilize the collected data and samples. The investigator must ensure that the person enrolling in the research study has had time to take a decision freely and has read and understood the information sheet and consent form. The investigator will inform the person enrolling in the research study that consent is an ongoing process and that he /she is entitled to withdraw from the study at any time, without any bias, discrimination or impact on their medical care.

#### Confidentiality

Subjects will not be identified by their names, but only by their assigned subject code. Raw and curated data files and databases will be stored on the password-protected servers of each institute behind a secure firewall. File read/write permissions will be restricted to members of the project team and other partners. Project coordinators and data managers will be responsible for data integrity. The Information Security service will be responsible for data security. Data access will be authorized to requestors after evaluation of a data access request that includes the objectives and methods of the requestor’s project, and a signed data access agreement. This agreement lists the conditions that govern access to the data, relative to intellectual property, ethical considerations, and data security.

## Discussion

The One Health approach is increasingly recognized as essential in addressing zoonotic and environmentally linked diseases, including vector- and water-borne infections [44]. By integrating human, animal and environmental health, One Health allows for a more holistic understanding of disease emergence and transmission within complex ecological and social systems. This systems-level perspective supports the identification of risk pathways and informs more effective and sustainable prevention and control strategies [45].

Operationalizing One Health involves more than collecting data across human, animal, and environmental compartments; it requires coordinated, cross-sectoral systems that translate evidence into action. This includes joint surveillance, shared data interpretation, and integrated responses across institutions and disciplines. Activities such as molecular surveillance in domestic and wild animals, genomic comparisons of pathogens across hosts, and environmental sampling incorporating climatic and hydrological variables (e.g., rainfall, flooding, land use) are critical technical components of this framework. Their values lie in how they are connected to decision-making processes and public health intervention strategies that benefit all three domains – for example, informing targeted vaccination in humans or animals, improving waste and water management or enhancing vector control at community and ecosystem levels. This integrative approach is particularly relevant for zoonotic, vector-borne, and waterborne diseases, where meteorological and ecological factors directly shape transmission dynamics and where effective response depends on collaboration across sectors.

Expanding surveillance across human, animal and environmental compartments introduces both methodological and operational challenges. Study designs must be adapted to the specific characteristics of each domain while ensuring coherence and representativeness across compartments to effectively address complex research questions [45]. One Health implementation also requires sustained multisectoral collaboration[46], often involving professionals from distinct disciplines who may use different technical languages, follow varied institutional mandates[47], and work across geographically dispersed or resource-limited settings [48].

Despite these challenges, when rigorously applied, One Health approaches can provide critical insights into transmission pathways and significantly enhance early detection systems for emerging and re-emerging infectious diseases. The COVID-19 pandemic – with its likely zoonotic origin, rapid global spread, and the urgent need for coordinated multisectoral response – brought renewed global attention to the importance of integrated surveillance and preparedness. It exposed the vulnerabilities of fragmented systems and underscored the necessity of Early Warning mechanisms that span human, animal, and environmental health sectors. In Cambodia, where surveillance infrastructure and diagnostic capacity remain limited, an integrated approach that bridges these gaps could enable more timely and effective responses to health threats at the human–animal–environment interface. While the value of One Health is widely acknowledged, questions persist regarding its cost-effectiveness and long-term sustainability[49]. Several studies emphasize the need for robust, quantitative evaluation of the impact of One Health, including cost-benefit analyses, to demonstrate value of these approaches over traditional siloed systems [50]. In regions such as Southeast Asia, limited laboratory capacity and human resources may hinder the sustainability and impact of One Health initiatives once external funding ends [51].

Addressing these gaps requires sustained investment in capacity building that is locally grounded, technically relevant, and co-developed with national authorities across all relevant ministries and sectors. Th establishment of the Interministerial Coordinating Committee on One Health (IMCC-OH) in Cambodia in August 2023 – bringing together the Ministry of Health, the Ministry of Agriculture, Forestry and Fisheries, and the Ministry of Environment – marks a significant step toward formalizing multisectoral coordination and provides a structured platform for advancing multisectoral approaches.

This study is designed to contribute directly to these national efforts by providing a field-based example of integrated surveillance and data collection across human, animal, and environmental domains. By demonstrating the practical implementation of One Health at the community level, the protocol aims to inform future research and guide policy development. Moreover, it addresses the challenge of shifting from conventional single-sector methods toward operationalized, multi-domain One Health systems.

## Contribution

This study will generate integrated data on ecological, veterinary and public health drivers of infectious disease transmission in Cambodia, providing a solid evidence base to inform public health strategies tailored to local realities and to strengthen surveillance and epidemic response capacities. The longitudinal, cross-sectorial dataset will also yield tools and protocols that could support sub-national strategies for community-based early-warning systems – a critical need given Cambodia’s currently fragmented surveillance landscape and ongoing risks of disease emergence[52].

Identifying human and/or animal populations at risk will allow the design of targeted and cost-effective awareness or vaccination campaigns (where vaccines are available). Recognizing at-risk practices across compartments will support the implementation of effective barrier measures adapted to seasonal changes and ecological variations. Furthermore, linking environmental data to pathogen circulation will help to inform preventive environmental measures – such as improving access to safe water, enhancing waste management, protecting water sources, and mitigating environmental degradation caused by human activities.

By mapping disease burden across compartments, this research can help prioritize pathogens of concern and guide resource allocation toward the most impactful interventions [46]. It also shed light on neglected diseases that are ecologically significant but under-recognized, especially those linked to animal reservoirs or environmental exposure, and thus particularly well suited for One Health approaches [53].

Ultimately, this study aims to move beyond knowledge generation to provide actionable evidence for decision-makers. By bridging research, surveillance, and policy, it offers a practical model for implementing One Health in rural communities, with lessons potentially transferable to other regions. The findings are intended to strengthen multisectoral collaboration, inform national One Health strategies, and enhance preparedness for emerging infectious diseases.

## Limitations

A potential methodological limitation could arise from a temporal mismatch across compartments if, due to logistical constraints, there is a delay between the collection of human, animal or environmental samples. Such a gap could limit the ability to capture truly synchronous infection events and may introduce bias in interpreting cross-compartment transmission patterns. In addition, differences in the nature of the indicators measured add another layer of complexity: in humans, serological testing reflects cumulative exposure to pathogens over an extended period, capturing both recent and past infections, whereas molecular detection in animals, vectors, and environmental samples provides only a snapshot of current or very recent infections or contamination. Together, these differences in timing and biological signal require cautious interpretation and underscore the need for integrated modeling approaches and triangulation with ecological, climatic, and behavioral data to strengthen inferences on transmission dynamics.

Diagnostic limitations may also affect data precision. For instance, the serological tools employed exhibit reduced sensitivity and specificity for certain pathogens, especially due to cross-reactivity between antigenically related flaviviruses like DENV, ZIKV and JEV. Such issues could lead to over-or under-estimation of true exposure. To mitigate this, confirmatory testing such as neutralization assays will be performed and results further refined using mathematical models to adjust for cross-reactivity and other confounding factors.

Biases may also arise from the self-reported nature of KAP data as well as infection history recall. Social desirability bias, recall errors, misreporting, and limited health literacy in some households may affect the accuracy of responses. The absence of direct behavioral observation adds further uncertainty to interpretations regarding risk practices and exposure pathways.

Finally, unmeasured environmental and external factors, including seasonal fluctuations, extreme weather events, and *force majeure* disruptions such as natural disasters or sociopolitical instability, may influence transmission risk across time and space, further complicating temporal comparisons between data collection rounds.

Despite these constraints, the integrated and multi-compartmental design of the study is a major strength. By combining serological, molecular, environmental and behavioral data within in an integrated framework, the study protocol offers a uniquely holistic and comprehensive perspective on zoonotic transmission dynamics in rural Cambodia. While immediate generalizability may be limited, the results are expected to provide critical insights into pathogen ecology, human–animal–environment interactions, and local drivers of infection risk, forming a robust foundation for targeted, context-specific surveillance strategies, interventions, and future research in similar ecological and socio-economic settings.

## Conclusion

This protocol outlines an ambitious and integrated One Health study designed to capture the complexity of zoonotic and environmentally driven disease transmission in rural Cambodia. By bridging human, animal, and environmental data across space and time, and accounting for hydrological variations and seasonality, the study seeks to move beyond traditional surveillance to generate actionable insights for targeted interventions and risk reduction. The design aims to be both scientifically robust and policy-relevant, aligning with Cambodia’s national One Health agenda while offering a framework that could be adapted to other regions facing similar ecological and institutional challenges.

## Author contributions

**Conceptualization**: Anne-Laure Bañuls, Sébastien Boyer, Sokleaph Cheng, Simon Cauchemez, Veasna Duong, Claude Flamand, Flavie Goutard, Hélène Guis, Vincent Herbreteau, Erik Karlsson, Sowath Ly

**Methodology**: Andrea Antoniolli, Juliana Aizawa Porto de Abreu, Anne-Laure Bañuls, Sébastien Boyer, Sokleaph Cheng, Tineke Cantaert, Veasna Duong, Claude Flamand, Hélène Guis, Bertrand Guillard, Vincent Herbreteau, Mallorie Hide, Erik Karlsson, Kunthy Nguon, Janin Nouhin, Sopheak Sorn

**Investigation**: Andrea Antoniolli, Juliana Aizawa Porto de Abreu, Sébastien Boyer, Sokleaph Cheng, Sopheak Chhay, Veasna Duong, Claude Flamand, Hélène Guis, Savatey Hak, Mallorie Hide, Vibol Hul, Phanit Kong, Sowath Ly, Kimsear Nov, Kunthy Nguon, Kimyeun Oeurn, Téphanie Sieng, Sopheak Sorn

**Formal analysis**: Andrea Antoniolli, Sébastien Boyer, Simon Cauchemez, Claude Flamand, Hélène Guis, Vincent Herbreteau

**Data curation**: Andrea Antoniolli, Juliana Aizawa Porto de Abreu, Phanit Kong, Kimsear Nov, Kimyeun Oeurn, Sopheak Sorn

**Resources**: Tineke Cantaert, Flavie Goutard, Bertrand Guillard, Erik Karlsson, Sidonn Krang

**Visualization**: Andrea Antoniolli

**Supervision**: Anne-Laure Bañuls, Sébastien Boyer, Claude Flamand, Flavie Goutard, Bertrand Guillard, Vincent Herbreteau, Mallorie Hide, Erik Karlsson, Sowath Ly

**Funding acquisition**: Anne-Laure Bañuls, Simon Cauchemez, Claude Flamand

**Writing – Original Draft Preparation**: Andrea Antoniolli, Sébastien Boyer, Claude Flamand, Mallorie Hide

**Writing – Review & Editing**: All authors

## Data availability statement

No datasets were generated or analysed for the current manuscript as this article describes a study protocol. Data generated during the AFRICAM study will be made publicly available in accordance with institutional policies and applicable ethical and legal regulations.

## Data Availability

All data produced in the present work are contained in the manuscript.

## Acknowledgments

We thank the Institut de Recherche pour le Développement (IRD), the Centre de Coopération Internationale en Recherche Agronomique pour le Développement (CIRAD), the Institute of Technology of Cambodia (ITC), and the Institut Pasteur du Cambodge (IPC) for their technical and scientific support. We are grateful to the Battambang provincial authorities for facilitating field access and coordination, to the Interministerial Coordinating Committee on One Health (IMCC-OH) for cross-sectoral guidance, and to the Communicable Disease Control Department of the Ministry of Health (CDC-MoH) for its collaboration.

We extend special thanks to the Ministry of Agriculture, Forestry and Fisheries (MAFF)—in particular the General Directorate of Animal Health and Production (GDAPH) for livestock and domestic-animal work, and the Forestry Administration for wildlife activities—for their on-site supervision, attendance, and active participation in all animal-sector field investigations, ensuring that procedures were conducted by qualified personnel in accordance with national practice and animal-welfare standards. We also acknowledge the Ministry of Environment (MoE) for its support.

Finally, we thank Battambang Hospital, village chiefs, and community representatives for logistical assistance, and the residents of the study sites for their active participation and support.

## References

[1] Jones KE, Patel NG, Levy MA, Storeygard A, Balk D, Gittleman JL, et al. Global trends in emerging infectious diseases. Nature 2008;451:990–3. 10.1038/nature06536.

[2] Hierink F, Okiro EA, Flahault A, Ray N. The winding road to health: A systematic scoping review on the effect of geographical accessibility to health care on infectious diseases in low-and middle-income countries. PLoS ONE 2021;16:e0244921. 10.1371/journal.pone.0244921.

[3] Morand S, Blasdell K, Bordes F, Buchy P, Carcy B, Chaisiri K, et al. Changing landscapes of Southeast Asia and rodent-borne diseases: decreased diversity but increased transmission risks. Ecol Appl 2019;29:e01886. 10.1002/eap.1886.

[4] Kazama S, Aizawa T, Watanabe T, Ranjan P, Gunawardhana L, Amano A. A quantitative risk assessment of waterborne infectious disease in the inundation area of a tropical monsoon region. Sustain Sci 2012;7:45–54. 10.1007/s11625-011-0141-5.

[5] Watts N, Amann M, Ayeb-Karlsson S, Belesova K, Bouley T, Boykoff M, et al. The *Lancet* Countdown on health and climate change: from 25 years of inaction to a global transformation for public health. The Lancet 2018;391:581–630. 10.1016/S0140-6736(17)32464-9.

[6] Horm SV, Tarantola A, Rith S, Ly S, Gambaretti J, Duong V, et al. Intense circulation of A/H5N1 and other avian influenza viruses in Cambodian live-bird markets with serological evidence of sub-clinical human infections. Emerg Microbes Infect 2016;5:e70. 10.1038/emi.2016.69.

[7] Edwards KM, Siegers JY, Wei X, Aziz A, Deng Y-M, Yann S, et al. Detection of Clade 2.3.4.4b Avian Influenza A(H5N8) Virus in Cambodia, 2021 - Volume 29, Number 1—January 2023 - Emerging Infectious Diseases journal - CDC n.d. 10.3201/eid2901.220934.

[8] Horwood PF, Horm SV, Yann S, Tok S, Chan M, Suttie A, et al. Aerosol exposure of live bird market workers to viable influenza A/H5N1 and A/H9N2 viruses, Cambodia. Zoonoses Public Health 2023;70:171–5. 10.1111/zph.13009.

[9] Hem S, Ly S, Votsi I, Vogt F, Asgari N, Buchy P, et al. Estimating the Burden of Leptospirosis among Febrile Subjects Aged below 20 Years in Kampong Cham Communities, Cambodia, 2007-2009. PloS One 2016;11:e0151555. 10.1371/journal.pone.0151555.

[10] Berlioz-Arthaud A, Guillard B, Goarant C, Hem S. [Hospital-based active surveillance of human leptospirosis in Cambodia]. Bull Soc Pathol Exot 1990 2010;103:111–8. 10.1007/s13149-010-0043-2.

[11] Cousien A, Ledien J, Souv K, Leang R, Huy R, Fontenille D, et al. Predicting Dengue Outbreaks in Cambodia. Emerg Infect Dis 2019;25:2281–3. 10.3201/eid2512.181193.

[12] Teurlai M, Huy R, Cazelles B, Duboz R, Baehr C, Vong S. Can human movements explain heterogeneous propagation of dengue fever in Cambodia? PLoS Negl Trop Dis 2012;6:e1957. 10.1371/journal.pntd.0001957.

[13] Manning JE, Oliveira F, Parker DM, Amaratunga C, Kong D, Man S, et al. The PAGODAS protocol: pediatric assessment group of dengue and Aedes saliva protocol to investigate vector-borne determinants of Aedes-transmitted arboviral infections in Cambodia. Parasit Vectors 2018;11:664. 10.1186/s13071-018-3224-7.

[14] Auerswald H, Ruget A-S, Ladreyt H, In S, Mao S, Sorn S, et al. Serological Evidence for Japanese Encephalitis and West Nile Virus Infections in Domestic Birds in Cambodia. Front Vet Sci 2020;7:15. 10.3389/fvets.2020.00015.

[15] Di Francesco J, Choeung R, Peng B, Pring L, Pang S, Duboz R, et al. Comparison of the dynamics of Japanese encephalitis virus circulation in sentinel pigs between a rural and a peri-urban setting in Cambodia. PLoS Negl Trop Dis 2018;12:e0006644. 10.1371/journal.pntd.0006644.

[16] Pommier JD, Gorman C, Crabol Y, Bleakley K, Sothy H, Santy K, et al. Childhood encephalitis in the Greater Mekong region (the SouthEast Asia Encephalitis Project): a multicentre prospective study. Lancet Glob Health 2022;10:e989–1002. 10.1016/S2214-109X(22)00174-7.

[17] Yek C, Li Y, Pacheco AR, Lon C, Duong V, Dussart P, et al. National dengue surveillance, Cambodia 2002-2020. Bull World Health Organ 2023;101:605–16. 10.2471/BLT.23.289713.

[18] Champagne C, Paul R, Ly S, Duong V, Leang R, Cazelles B. Dengue modeling in rural Cambodia: Statistical performance versus epidemiological relevance. Epidemics 2019;26:43–57. 10.1016/j.epidem.2018.08.004.

[19] Ly S, Fortas C, Duong V, Benmarhnia T, Sakuntabhai A, Paul R, et al. Asymptomatic Dengue Virus Infections, Cambodia, 2012-2013. Emerg Infect Dis 2019;25:1354–62. 10.3201/eid2507.181794.

[20] Doeurk B, Marcombe S, Maquart P-O, Boyer S. Review of dengue vectors in Cambodia: distribution, bionomics, vector competence, control and insecticide resistance. Parasit Vectors 2024;17:424. 10.1186/s13071-024-06481-5.

[21] Census of Agriculture Cambodia 2023. National Institute of Statistics (NIS), Ministry of Planning (MoP), Ministry of Agriculture, Forestry and Fisheries (MAFF), FAO Cambodia; 2024.

[22] Coker RJ, Hunter BM, Rudge JW, Liverani M, Hanvoravongchai P. Emerging infectious diseases in southeast Asia: regional challenges to control. Lancet Lond Engl 2011;377:599–609. 10.1016/S0140-6736(10)62004-1.

[23] Elnaiem A, Mohamed-Ahmed O, Zumla A, Mecaskey J, Charron N, Abakar MF, et al. Global and regional governance of One Health and implications for global health security. The Lancet 2023;401:688–704. 10.1016/S0140-6736(22)01597-5.

[24] Mwangi W, Figueiredo P de, Criscitiello MF. One Health: Addressing Global Challenges at the Nexus of Human, Animal, and Environmental Health. PLOS Pathog 2016;12:e1005731. 10.1371/journal.ppat.1005731.

[25] Panel (OHHLEP) OHH-LE, Adisasmito WB, Almuhairi S, Behravesh CB, Bilivogui P, Bukachi SA, et al. One Health: A new definition for a sustainable and healthy future. PLOS Pathog 2022;18:e1010537. 10.1371/journal.ppat.1010537.

[26] Antoniolli A, Guis H, Picardeau M, Goarant C, Flamand C. One Health Field Approach Applied to Leptospirosis: A Systematic Review and Meta-Analysis Across Humans, Animals and the Environment. Open Forum Infect Dis 2025;12:ofae757. 10.1093/ofid/ofae757.

[27] Khan MS, Rothman-Ostrow P, Spencer J, Hasan N, Sabirovic M, Rahman-Shepherd A, et al. The growth and strategic functioning of One Health networks: a systematic analysis. Lancet Planet Health 2018;2:e264–73. 10.1016/S2542-5196(18)30084-6.

[28] Gebreyes WA, Dupouy-Camet J, Newport MJ, Oliveira CJB, Schlesinger LS, Saif YM, et al. The global one health paradigm: challenges and opportunities for tackling infectious diseases at the human, animal, and environment interface in low-resource settings. PLoS Negl Trop Dis 2014;8:e3257. 10.1371/journal.pntd.0003257.

[29] W.b K, C S. Introduction - Is One Health delivering results 2014;33(2):375. 10.20506/rst.33.2.2301.

[30] Diallo AOI, Chevalier V, Cappelle J, Duong V, Fontenille D, Duboz R. How much does direct transmission between pigs contribute to Japanese Encephalitis virus circulation? A modelling approach in Cambodia. PLOS ONE 2018;13:e0201209. 10.1371/journal.pone.0201209.

[31] Lwanga S, Lemeshow S, World Health Organization (WHO). Sample Size Determination in Health Studies: A Practical Manual. XF2006303260 1991;15. 10.2307/2290547.

[32] National Research Council (US) Committee for the Update of the Guide for theCare and Use of Laboratory Animals. Guide for the Care and Use of Laboratory Animals. 8th ed. Washington (DC): National Academies Press (US); 2011.

[33] Rozier H, Gloaguen P, Septier F, Boyer S. Evaluating Experimental Design to Sample Mosquitoes 2025. 10.21203/rs.3.rs-6704646/v1.

[34] Manning JE, Chea S, Parker DM, Bohl JA, Lay S, Mateja A, et al. Development of Inapparent Dengue Associated With Increased Antibody Levels to Aedes aegypti Salivary Proteins: A Longitudinal Dengue Cohort in Cambodia. J Infect Dis 2022;226:1327–37. 10.1093/infdis/jiab541.

[35] Bailly S, Rousset D, Fritzell C, Hozé N, Ben Achour S, Berthelot L, et al. Spatial Distribution and Burden of Emerging Arboviruses in French Guiana. Viruses 2021;13:1299. 10.3390/v13071299.

[36] Beck C, Desprès P, Paulous S, Vanhomwegen J, Lowenski S, Nowotny N, et al. A High-Performance Multiplex Immunoassay for Serodiagnosis of Flavivirus-Associated Neurological Diseases in Horses. BioMed Res Int 2015;2015:678084. 10.1155/2015/678084.

[37] Flamand C, Bailly S, Fritzell C, Berthelot L, Vanhomwegen J, Salje H, et al. Impact of Zika Virus Emergence in French Guiana: A Large General Population Seroprevalence Survey. J Infect Dis 2019;220:1915–25. 10.1093/infdis/jiz396.

[38] Hozé N, Salje H, Rousset D, Fritzell C, Vanhomwegen J, Bailly S, et al. Reconstructing Mayaro virus circulation in French Guiana shows frequent spillovers. Nat Commun 2020;11:2842. 10.1038/s41467-020-16516-x.

[39] Fontaine A, Diouf I, Bakkali N, Missé D, Pagès F, Fusai T, et al. Implication of haematophagous arthropod salivary proteins in host-vector interactions. Parasit Vectors 2011;4:187. 10.1186/1756-3305-4-187.

[40] Elanga Ndille E, Doucoure S, Damien G, Mouchet F, Drame PM, Cornelie S, et al. First Attempt To Validate Human IgG Antibody Response to Nterm-34kDa Salivary Peptide as Biomarker for Evaluating Exposure to Aedes aegypti Bites. PLoS Negl Trop Dis 2012;6. 10.1371/journal.pntd.0001905.

[41] Elanga Ndille E, Doucoure S, Poinsignon A, Mouchet F, Cornelie S, D’Ortenzio E, et al. Human IgG Antibody Response to Aedes Nterm-34kDa Salivary Peptide, an Epidemiological Tool to Assess Vector Control in Chikungunya and Dengue Transmission Area. PLoS Negl Trop Dis 2016;10:e0005109. 10.1371/journal.pntd.0005109.

[42] Sagna AB, Yobo MC, Elanga Ndille E, Remoue F. New Immuno-Epidemiological Biomarker of Human Exposure to Aedes Vector Bites: From Concept to Applications. Trop Med Infect Dis 2018;3. 10.3390/tropicalmed3030080.

[43] Yobo CM, Sadia-Kacou CAM, Adja MA, Elanga-Ndille E, Sagna AB, Guindo-Coulibaly N, et al. Evaluation of Human Exposure to Aedes Bites in Rubber and Palm Cultivations Using an Immunoepidemiological Biomarker. BioMed Res Int 2018;2018:3572696. 10.1155/2018/3572696.

[44] Miao L, Li H, Ding W, Lu S, Pan S, Guo X, et al. Research Priorities on One Health: A Bibliometric Analysis. Front Public Health 2022;10:889854. 10.3389/fpubh.2022.889854.

[45] Promoting the science of One Health. Nat Commun 2023;14:4735. 10.1038/s41467-023-40293-y.

[46] J.r S. Importance of a One Health approach in advancing global health security and the Sustainable Development Goals 2019;38(1):145. 10.20506/rst.38.1.2949.

[47] Antoniolli A, Flamand C. Integrating One Health: Beyond buzzwords and silos. One Health 2025;21:101174. 10.1016/j.onehlt.2025.101174.

[48] Barnett T, Pfeiffer DU, Ahasanul Hoque M, Giasuddin M, Flora MS, Biswas PK, et al. Practising co-production and interdisciplinarity: Challenges and implications for one health research. Prev Vet Med 2020;177:104949. 10.1016/j.prevetmed.2020.104949.

[49] Winkler AS, Brux CM, Carabin H, das Neves CG, Häsler B, Zinsstag J, et al. The Lancet One Health Commission: harnessing our interconnectedness for equitable, sustainable, and healthy socioecological systems. Lancet Lond Engl 2025:S0140–6736(25)00627-0. 10.1016/S0140-6736(25)00627-0.

[50] Baum SE, Machalaba C, Daszak P, Salerno RH, Karesh WB. Evaluating one health: Are we demonstrating effectiveness? One Health 2016;3:5–10. 10.1016/j.onehlt.2016.10.004.

[51] Dahal R, Upadhyay A, Ewald B. One Health in South Asia and its challenges in implementation from stakeholder perspective. Vet Rec 2017;181:626. 10.1136/vr.104189.

[52] Avian Influenza A (H5N1) Outbreak 2024 in Cambodia: Worries Over the Possible Spread of the Virus to Other Asian Nations and the Strategic Outlook for its Control n.d. 10.1177/11786302241246453.

[53] Peterson JK, Bakuza J, Standley CJ. One Health and Neglected Tropical Diseases—Multisectoral Solutions to Endemic Challenges. Trop Med Infect Dis 2020;6:4. 10.3390/tropicalmed6010004.

